# Outdoor Nighttime Light Exposure (Light Pollution) is Associated with Alzheimer’s Disease

**DOI:** 10.1101/2024.02.14.24302831

**Authors:** Robin M Voigt, Bichun Ouyang, Ali Keshavarzian

## Abstract

Alzheimer’s disease (AD) prevalence has increased in the last century which can be attributed to increased lifespan, but environment is also important. This study evaluated the relationship between outdoor nighttime light exposure and AD prevalence in the United States. Higher outdoor nighttime light was associated with higher prevalence of AD. While atrial fibrillation, diabetes, hyperlipidemia, hypertension, and stroke were associated more strongly with AD prevalence than nighttime light intensity, nighttime light was more strongly associated with AD prevalence than alcohol abuse, chronic kidney disease, depression, heart failure, and obesity. Startlingly, nighttime light exposure more strongly associated with AD prevalence in those under the age of 65 than any other disease factor examined. These data indicate a need to investigate how nighttime light exposure influences AD pathogenesis.

**One-Sentence Summary:** There is a positive association between Alzheimer’s disease prevalence and average outdoor nighttime light intensity in the United States.

## Introduction

Throughout most of human history fire was used as the source of light (e.g., wood, tallow, wax, or oil), gas lighting emerged at the end of the 18^th^ century, and electric lighting was developed in the mid-19^th^ century, and by the early 1900s most homes in the United States had electric lighting (Bowers 1998; Brox 2010; Schivelbusch 1995; Boyle 2019). Nowadays artificial lights ubiquitously illuminate our indoor and outdoor spaces. Artificial outdoor lights provide safety, convenience, and aesthetics (e.g., deter crime, illuminate roadways, highlight landscaping), but excessive artificial light at night is called light pollution (e.g., poorly shielded, overly bright lighting fixtures). Today, most people living in urban and suburban areas are unable to see natural celestial light due to light pollution and as much as 80% of the global population experience light pollution. Although artificial light at night is considered by most to be harmless or even beneficial (e.g., safety), light pollution has detrimental ecological, behavioral, biological, and health consequences (Bedrosian and Nelson 2013).

Exposure to light at night is associated with numerous detrimental health effects including sleep disruption, obesity, depression, anxiety, memory dysfunction, atherosclerosis, and cancer (Bozejko, Tarski, and Malodobra-Mazur 2023) but little is known about the impact of light pollution on Alzheimer’s disease (AD). AD is the most common neurodegenerative disorder and accounts for 60-80% of dementia cases (Kapasi, DeCarli, and Schneider 2017; Brenowitz et al. 2017) and it is estimated that 10.8% of adults over the age of 65 have AD (Rajan et al. 2021). Incidence and prevalence of AD have increased in recent decades which parallels the increase in light pollution. Evidence suggests that exposure to light at night may promote neurodegeneration, neuroinflammation, dementia, and AD (Kim et al. 2018; Delorme et al. 2022; Walker et al. 2020; Chen et al. 2022; Mazzoleni et al. 2023; Romeo et al. 2013), but this possibility has not been carefully examined.

This study evaluated the relationship between AD prevalence and average nighttime light intensity (light pollution) in the United States (i.e., the lower 48 states) using data from the Centers for Medicare & Medicaid Services (Chronic Conditions), the Centers for Disease Control and Prevention (CDC, Behavioral Risk Factor Surveillance System (BRFSS)), and satellite-acquired light pollution data (VIIRS) (Details in Supplementary Materials).

## Results

### Average Nighttime Light Intensity is Associated with Higher Prevalence of AD

AD prevalence was acquired from Medicare data (**Fig 1a**) and average nighttime light intensity was generated from satellite acquired data (**Fig 1b**) and the data were averaged for years 2012-2018. States were ranked according to average nighttime light intensity and were divided into five groups representing those with the lowest average light intensity (i.e., 1/5, darkest) to the highest average light intensity (i.e., 5/5, brightest) (**Fig 1c**). Analysis revealed a statistically significant difference in AD prevalence between groups (F(4,43)=13.500, p<0.001). Multiple comparisons testing found statistical differences between states with the darkest average nighttime light intensity and states with brightest average nighttime light intensity (**Fig 1d**). Pearson correlation analysis similarly demonstrated a relationship between light intensity and AD prevalence (r(46)=0.557, p<0.001, **Fig 1e**). This relationship also existed when examining those above the age of 65 (ANOVA: F(4,43)=14.560, p<0.001; Correlation: r(46)=0.490, p<0.001) and those under the age of 65 (ANOVA: F(4,43)=6.950, p<0.001; Correlation: r(46)=0.550, p<0.001) (**Fig S1**). Each year was also assessed individually, and the same positive relationship was observed between nighttime light intensity and AD prevalence for each year independently (**Table S1**).

**Fig. 1.**
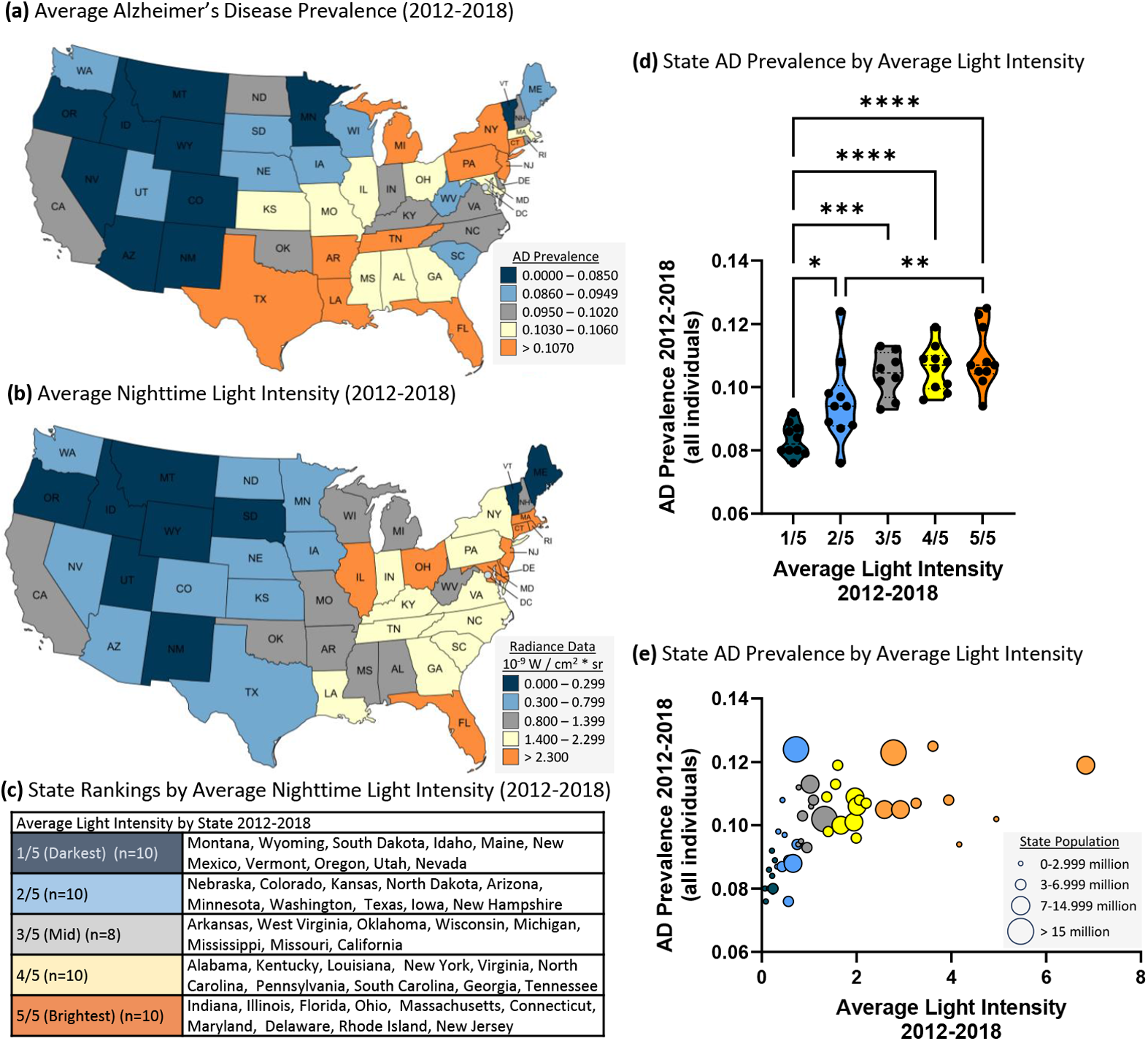
Higher average nighttime light intensity is associated with higher AD prevalence (2012-2018 average). **(a)** AD prevalence by state. **(b)** Average nighttime light intensity by state. **(c)** Average nighttime light intensity state rankings. 1/5 = darkest average nighttime light intensity through 5/5 = brightest average nighttime light intensity. **(d)** AD prevalence by state average nighttime light intensity rankings. ANOVA revealed a statistically significant difference in AD prevalence between groups (F(4,43)=13.50, p<0.0001). Multiple comparisons testing found that the states with the lowest average light intensity had statistically significant lower AD prevalence than states with higher average nighttime light intensity: 1/5 vs. 2/5 (p=0.031), 1/5 vs. 3/5 (p<0.001), 1/5 vs. 4/5 (p<0.001), 1/5 vs. 5/5 (p<0.001), and 2/5 vs. 5/5 (p=0.010). *p<0.05, **p<0.01, ***p<0.001, ****p<0.0001. **(e)** Pearson correlation analysis between AD prevalence and nighttime light intensity revealed a positive relationship (r(46)=0.557. p<0.001). The circle size indicates state population.

A linear mixed model was applied to examine the overall effect of light intensity on AD prevalence (from 2012 to 2018) taking account of within-state correlation due to repeated measures. When examining all individuals, the overall regression was statistically significant (Chi-square=1081.9, df=1, p<0.0001). Average nighttime light intensity was significantly associated with AD prevalence (estimate=0.283, SE=0.102, p=0.006, **Table 1**). This effect was also observed in those over the age of 65, under the age of 65, in both men and women, and in all races examined except Asian Pacific Islander (**Table 1**).

**Table 1.**
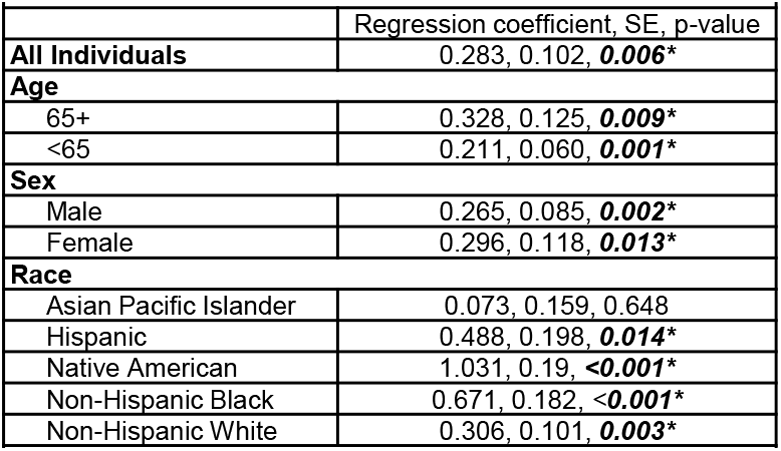
There is a significant relationship between AD prevalence and average nighttime light intensity from 2012-2018 across age, sex, and race / ethnicity. Linear mixed effects model data are provided for all individuals, age, sex, and race. AD prevalence was significantly associated with average nighttime light intensity in all groups except Asian Pacific Islander.

Co-variates known (or proposed to be) risk factors for AD including alcohol abuse, atrial fibrillation, chronic kidney disease, depression, diabetes, heart failure, hyperlipidemia, hypertension, obesity, and stroke were subsequently included in the model. Data for the covariates were obtained from Medicare Chronic Conditions data or from the CDC. When looking at all individuals, average nighttime light intensity was associated with AD prevalence even when accounting for alcohol abuse, chronic kidney disease, depression, heart failure, and obesity (**Table 2**). These data suggest that nighttime light intensity has a stronger influence on AD prevalence than these conditions. However, other covariates were more strongly associated with AD than light intensity including atrial fibrillation, diabetes, hyperlipidemia, hypertension, and stroke (**Table 2**) indicating that nighttime light exposure had a more subtle effect than these disease covariates. Similar results were obtained when examining men, women, and those over the age of 65 with each having a unique profile (**Table 2**). However, for those under the age of 65 average light intensity was associated with AD prevalence even when considering all the covariates (estimate=0.174, SE=0.064, p=0.007) (**Table 2**) suggesting that those under the age of 65 may be particularly sensitive to the effects of exposure to light at night.

**Table 2.** Average nighttime light intensity (2012-2018) is more strongly associated with AD prevalence than some known disease risk factors. Linear mixed effects model data are provided for ten chronic conditions that are established in the literature to impact risk of AD and/or AD incidence / prevalence. P Values are given for light pollution when considering the covariate. Non-significant p values indicate those covariates that have a stronger influence on AD than light pollution. CKD = chronic kidney disease

| Table 2. |  |  |  |  |  |
| --- | --- | --- | --- | --- | --- |
| Covariate | Regression coefficient, SE, p-value |  |  |  |  |
|  | All Individuals | Men | Woman | 65+ | <65 |
|  | 65+, <65, Men, Women | 65+ & <65 | 65+ & <65 | Men & Women | Men & Women |
| Light intensity (*Alcohol Abuse*) | 0.266, 0.104, **0.011\*** | 0.240, 0.087, **0.006\*** | 0.292, 0.119, **0.015\*** | 0.323, 0.126, **0.011\*** | 0.202, 0.061, **0.001\*** |
| Light intensity (*Atrial Fibrillation*) | 0.176, 0.098, 0.074 | 0.177, 0.084, **0.036\*** | 0.180, 0.108, 0.098 | 0.135, 0.118, 0.253 | 0.209, 0.060, **0.001\*** |
| Light intensity (*CKD*) | 0.289, 0.089, **0.001\*** | 0.238, 0.073, **0.001\*** | 0.312, 0.103, **0.003\*** | 0.325, 0.109, **0.003\*** | 0.209, 0.058, **<0.001\*** |
| Light intensity (*Depression*) | 0.295, 0.099, **0.003\*** | 0.260, 0.083, **0.002\*** | 0.329, 0.115, **0.005\*** | 0.376, 0.117, **0.001\*** | 0.201, 0.061, **0.001\*** |
| Light intensity (*Diabetes*) | 0.076, 0.091, 0.404 | 0.085, 0.080, 0.287 | 0.065, 0.192, 0.527 | 0.018, 0.106, 0.862 | 0.170, 0.061, **0.005\*** |
| Light intensity (*Heart Failure*) | 0.222, 0.072, **0.002\*** | 0.195, 0.067, **0.004\*** | 0.238, 0.077, **0.002\*** | 0.205, 0.080, **0.011\*** | 0.190, 0.059, **0.002\*** |
| Light intensity (*Hyperlipidemia*) | 0.145, 0.102, 0.157 | 0.136, 0.086, 0.117 | 0.132, 0.117, 0.262 | 0.143, 0.125, 0.253 | 0.163, 0.064, **0.012\*** |
| Light intensity (*Hypertension*) | 0.063, 0.093, 0.500 | 0.102, 0.084, 0.227 | 0.024, 0.101, 0.809 | -0.037, 0.104, 0.719 | 0.174, 0.063, **0.006\*** |
| Light intensity (*Stroke*) | 0.155, 0.092, 0.092 | 0.195, 0.079, **0.015\*** | 0.149, 0.105, 0.155 | 0.122, 0.107, 0.254 | 0.187, 0.060, **0.002\*** |
| Light intensity (*Obesity*) | 0.284, 0.102, **0.006\*** | data not available | data not available | data not available | data not available |
| Light intensity (*All Covariates*) | -0.004, 0.085, 0.967 | -0.006, 0.075, 0.933 | 0.017, 0.093, 0.857 | -0.068, -0.088, 0.441 | 0.174, 0.064, **0.007\*** |

### County Analysis

Each state is heterogeneous including urban, suburban, and rural areas each with different exposures (e.g., dietary habits/patterns, activity level, air pollution, light pollution). Therefore, we examined the relationship between average nighttime light intensity and AD prevalence in counties since these are smaller in size and more homogeneous than the state level data. To do so, the largest city in each state was identified and the county in which this city resided was noted and average nighttime light intensity was determined for that county and compared to AD prevalence from Medicare Chronic Conditions data at the county level. Using this approach, data was retrieved for 45 counties and the District of Columbia. The data were subsequently clustered based on average nighttime light intensity representing those counties with the lowest average nighttime light intensity (i.e., 1/4, darkest) to the highest average nighttime light intensity (i.e., 4/4, brightest) (**Fig 2a**). Analysis revealed a statistically significant difference in AD prevalence between groups (F(3,42)=10.750, p<0.001). Multiple comparisons testing found that states with the darkest average nighttime light intensity were statistically different from states with brightest average nighttime light intensity (**Fig 2b**). Pearson correlation analysis similarly demonstrated a relationship between light intensity and AD prevalence (r(44)=0.599, p<0.001, **Fig 2c**). This relationship also existed when examining those over the age of 65 (ANOVA: F(3,42)=10.840, p<0.001; Correlation: r(44)=0.612, p<0.001) and those under the age of 65 (ANOVA: F(3,42)=8.424, p<0.001; Correlation: r(44)=0.514, p<0.001) (**Fig S2**).

**Fig. 2:**
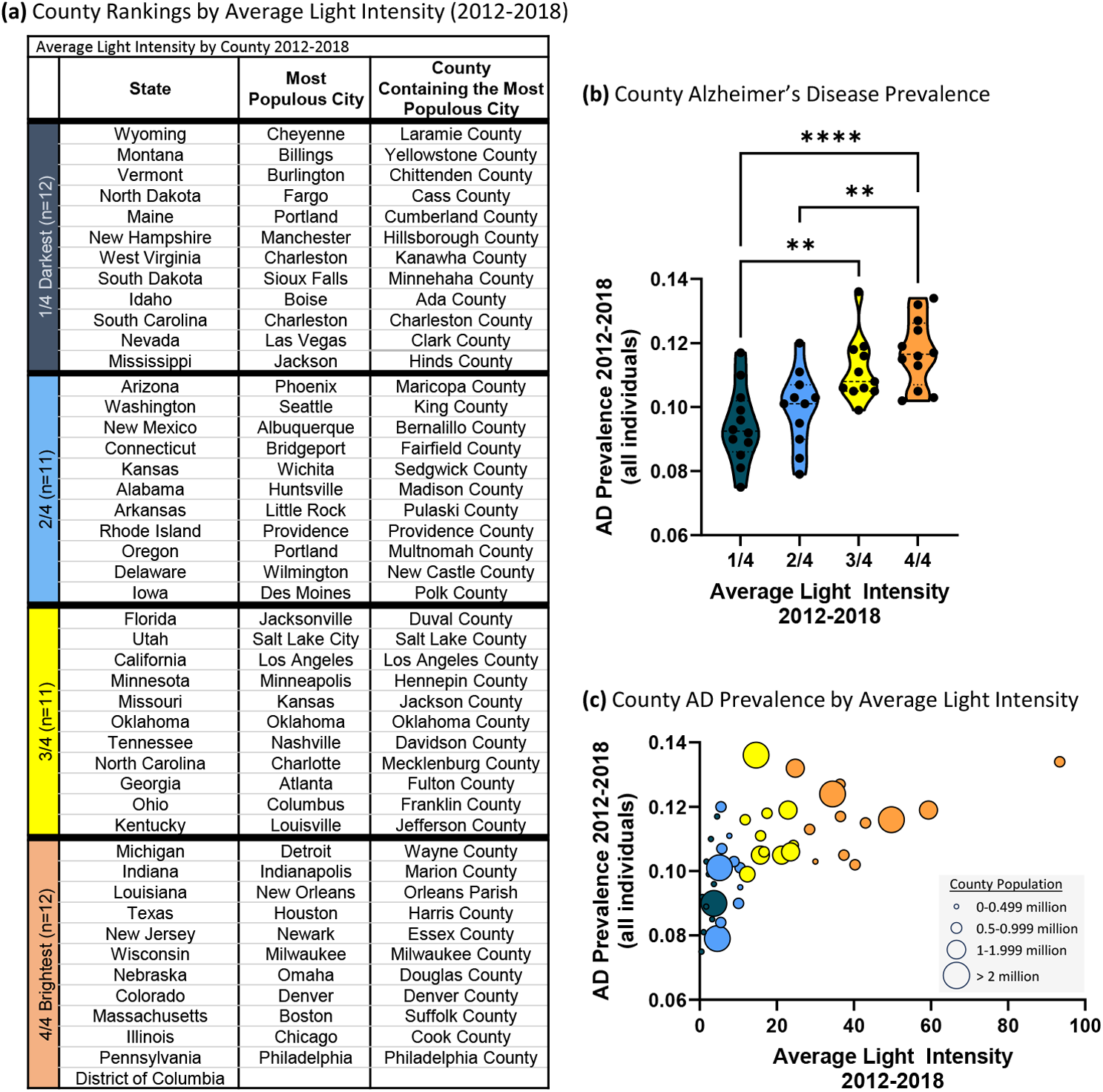
Higher county average nighttime light intensity is associated with higher county AD prevalence (2012-2018). **(a)** Average nighttime light intensity county rankings. AD prevalence by state average nighttime light intensity rankings. 1/5 = darkest average nighttime light intensity through 5/5 = brightest average nighttime light intensity. **(b)** AD prevalence by county average nighttime light intensity rankings. ANOVA: F(3,42)=10.750, p<0.001. Multiple comparisons testing revealed differences between groups with lower average light intensity and groups with higher average light intensity: 1/4 vs. 3/4 (p<0.01), 1/4 vs. 4/4 (p<0.001), 2/4 vs. 4/4 (p<0.01). **(c)** Pearson correlation analysis between AD prevalence and nighttime light intensity: r(44)=0.599, p<0.001. p<0.05, **p<0.01, ***p<0.001, ****p<0.0001. Circle size reflects state population.

A linear mixed effects model was applied to examine the overall effect of average light intensity on AD prevalence (from 2012 to 2018) considering within-county correlation due to repeated measures. When examining all individuals, average nighttime light intensity was significantly association with AD prevalence (estimate=0.040, SE=0.003, p<0.001) including in those over the age of 65 (estimate=0.052, SE=0.004, p<0.001) and under the age of 65 (estimate=0.019, SE=0.002, p<0.001). In summary, the results observed on the state level are recapitulated on the county level bolstering support for the positive association between AD prevalence and nighttime light exposure.

## Discussion

### Average Light Intensity is Associated with AD Prevalence

The analyses reveal that greater average nighttime light intensity (i.e., light pollution) was associated with higher AD prevalence. This was true for 2012-2018 average and each year examined individually, and in those over and under the age of 65 (i.e., 65+, <65), in both sexes, and in each race (except Asian Pacific Island which may be related to power). This finding was observed when examining data on the state level as well as on the county level.

Average nighttime light intensity was more strongly associated with AD prevalence than some diseases and conditions reported or suspected to be risk factors for AD including alcohol abuse (Andrews, Goate, and Anstey 2020; Anstey, Mack, and Cherbuin 2009; Wang et al. 2023; Nallapu et al. 2023), chronic kidney disease (Zhang et al. 2020; Tang et al. 2022), depression (Saiz-Vazquez et al. 2021), heart failure (Qiu et al. 2006; Cermakova et al. 2015), and obesity (Alford et al. 2018; Al-Kuraishy et al. 2023). However, other covariates were more strongly associated with AD than average nighttime light intensity including atrial fibrillation (Giannone et al. 2022), diabetes, hyperlipidemia, hypertension, and stoke. This finding is perhaps not surprising as the effects of nighttime light exposure would be expected to be more subtle than factors robustly associated with AD. However, in individuals under the age of 65, nighttime light intensity was associated with AD prevalence to a greater degree than all other disease risk factors, suggesting that younger individuals may be particularly sensitive to the detrimental effects of light at night. More research on how light at night may influence AD is needed.

### Nighttime Light Exposure: An Important Part of The Exposome

Dementia is not a modern phenomenon. Dementia is mentioned by Pythagoras, Hippocrates, Plato, and Shakespeare (Yang et al. 2016) and yet since the development of diagnostic criteria and AD incidence and prevalence have been reported, rates of AD have risen dramatically. This increase is attributed to factors like the aging population, emergence of lifestyle related diseases like obesity, high blood pressure, and diabetes, and a myriad of environmental factors related to urbanization and the industrial revolution. One such example is air pollution which is associated with cognitive decline (Franz et al. 2023), dementia (Gong et al. 2023), faster cognitive decline in those with AD (Lee et al. 2023), and incident neurodegenerative disease (Zhu et al. 2023). Exposure to light at night could similarly be impacting the development or progression of dementia and neurodegenerative disease.

To illustrate this possibility, we can examine Amish communities (who intentionally limit their use of modern technology and electricity and may consequently be exposed to less light at night). Amish communities have a lower prevalence of cognitive impairment in individuals of advanced age compared to the general population (Johnson et al. 1997; Holder and Warren 1998; Pericak-Vance et al. 1996). Numerous factors other than light exposure may contribute to this phenomenon (e.g., emphasis on family / community, traditional farming, genetics), but that being said, exposure of Drosophila to dim light during the dark period promotes neurodegeneration (Kim et al. 2018), and data from mice demonstrate that dim light during the dark period alters neuronal dendrites in AD-relevant brain regions (cortex, hippocampus) (Delorme et al. 2022), and long-term exposure to outdoor light at night is associated with increased risk of cognitive impairment in humans (Chen et al. 2022), and there is a positive correlation between light pollution and Parkinson’s disease (another neurodegenerative disease) (Romeo et al. 2013). Additional studies carefully evaluating indoor and outdoor light exposure and mechanistic evaluations are needed to fully understand the impact of nighttime light exposure and light pollution on AD.

The analyses in this report encompass years 2012 through 2018 during which time there was a trend for states to have a decrease in nighttime light (2012 = 100.000, 2018 = 96.890, p=0.054). The pandemic further decreased nighttime light exposure which was at the lowest level in 2020; however, despite the fact that 17 states have legislation designed to reduce light pollution nighttime light was the highest in 2022 (latest data available) (2012 = 100.000, 2022 = 103.500, p=0.007) (**Fig S3**). Given the association between nighttime light exposure and AD prevalence the increased nighttime light could potentially impact AD incidence and prevalence.

### Potential Mechanisms by Which Light Pollution May Promote Alzheimer’s Disease

There are multiple mechanisms by which exposure to light at night may influence AD development or progression. On the biochemical level, exposure of mice to dim light increases the production of the pro-inflammatory cytokine interleukin 1β (IL-1β) (Walker et al. 2020), which is a feature associated with AD. Additionally, exposure of mice to dim light during the dark period decreases levels of the neurotrophic factor brain derived neurotrophic factor (BDNF) in a brain region relevant for AD (i.e., the hippocampus) (Walker et al. 2020). This finding is intriguing as our group has demonstrated that low levels of BDNF are a feature that precede cognitive impairment (Voigt et al. 2021). Additionally, exposure to light at night may disrupt circadian rhythms (Raap, Pinxten, and Eens 2015) which is widely reported by our group and others to have detrimental effects on health. Indeed, changes in circadian rhythms often precede symptoms of AD in humans (Colwell 2021) and it is interesting to consider that exposure to light at night (via circadian disruption) may contribute to AD pathogenesis. Additionally, it is noteworthy that disruption of circadian rhythms is associated with increased risk of diseases that are risk factors for AD including obesity, diabetes, and depression, just to name a few (Fishbein, Knutson, and Zee 2021).

### Caveats

There are limitations associated with this study related to the curation of AD prevalence and light exposure data. First, the Medicare data are limited to those enrolled in Medicare Part A and Part B. Those with Medicare Advantage, only Part A, or only Part B are excluded; thus, the data presented in this report are not comprehensive (although Medicare data are reported to be reasonably reliable) (Grodstein et al. 2022). Second, the Medicare data report current residences of individuals which may not reflect life-long residence (important to understand causality). Third, this study evaluated AD prevalence and not incidence. Fourth, this study used state and county average light intensity data from satellites which does not represent indoor light exposures (e.g., televisions, computers, phones) which is important to consider to fully understand the impact of nighttime light on AD.

## Conclusion

Many (but not all) studies suggest that AD incidence has declined during the last decade (Matthews et al. 2016; Langa et al. 2017; Wolters et al. 2020; Rocca et al. 2011; Wu et al. 2017; Schrijvers et al. 2012; Satizabal et al. 2016; Cerasuolo et al. 2017; Derby et al. 2017; Ahmadi-Abhari et al. 2017; Sullivan et al. 2019; Matthews et al. 2013). This is thought to be attributed to better treatment of AD-associated risk factors (e.g., hypertension, diabetes), greater educational attainment, and perhaps greater awareness of factors that contribute to AD (e.g., diet) (Hudomiet, Hurd, and Rohwedder 2022). Data from this study suggest that reduction of these major factors may cause environmental factors like nighttime light exposure to have a substantial influence on AD pathogenesis in the future. Additionally, AD prevalence is expected to continue to increase due to increased life expectancy (Grodstein et al. 2022; Guerreiro and Bras 2015; Kawas et al. 2000; Fitten, Ortiz, and Ponton 2001; Matthews et al. 2019; Mayeda et al. 2016, 2017; Ajrouch, Zahodne, and Antonucci 2017) and it is of value to understand how exposure to light at night influences AD progression. While data from preclinical studies and the current study suggest exposure to light at night may influence AD, additional studies evaluating clinical and population health are needed.

## Methods

### Sex as a Biological Variable

De-identified data were obtained from Medicare reports of disease prevalence. All available data were included inclusive of both sexes.

### Alzheimer’s Disease Prevalence & Co-Variate Data Acquisition

#### Medicare Data

Medicare reports of disease prevalence from 2012-2018 were used. The data were obtained from the Office of Enterprise Data and Analytics, within the Centers for Medicare and Medicaid Services (CMS) which develops analytics examining chronic conditions among Medicare fee-for-service beneficiaries. Medicare is Federal a health insurance program for individuals over the age of 65, persons 65 years and under with certain disabilities, and individuals of any age with end-stage renal disease in the United States.

The data used in the chronic conditions report are based on CMS administrative enrollment and claims data for Medicare beneficiaries enrolled in the fee-for-service program. The data are available from the CMS Chronic Conditions Data Warehouse (CCW) (www.ccwdata.org). The chronic conditions public use files report prevalence in the Medicare beneficiary population limited to fee-for-service beneficiaries. Excluded populations include: (1) Medicare beneficiaries with any Medicare Advantage enrollment during the year and (2) beneficiaries who were enrolled in Part A only or Part B only. In 2018, exclusions accounted for approximately 44.9% of the total population. Beneficiaries who died during the year were included up to the date of death if other inclusion criteria were met.

The CMS CCW database includes pre-defined indicators for chronic conditions and mental health conditions (details on these conditions is available at www.ccwdata.org). A Medicare beneficiary is considered to have a chronic condition if a claim indicates that the beneficiary received a service or treatment for the specific condition. Chronic conditions are identified by diagnosis codes on the Medicare claims. Services prior to October 2015 used International Classification of Diseases version 9 (ICD-9-CM) codes, chronic conditions identified in or after October 2015 used version 10 (ICD-10-CM-December). Beneficiaries may have more than one chronic condition. Estimates of the prevalence of Chronic Conditions may vary from other sources, as estimates of Chronic Conditions will be influenced by the number and type of conditions that are used. Geographic variation in the prevalence estimates of chronic conditions can be affected by using diagnoses on administrative claims to infer the presence of a condition. Variability in coding diagnoses can lead to both the under and over diagnosis of specific conditions and affect estimates of chronic conditions.

Estimates are measures of overall magnitude of chronic conditions in the Medicare population for national, state, and county levels. Data are presented based upon the beneficiary’s residence, rather than where care was received. Prevalence estimates are not age or sex adjusted nor are they adjusted for beneficiary characteristics across geographic variability. There is evidence that regional variation in care is associated with the supply of health care resources, which can affect prevalence estimates; since in places where more healthcare resources are available, the likelihood that diagnoses will be identified may be increased. A recent assessment found that Medicare claims perform reasonably well in identifying dementia (Grodstein et al. 2022).

#### Center for Disease Control and Prevention Data

Obesity data were obtained from the Center for Disease Control and Prevention (CDC). In 1984 the CDC initiated the state-based Behavioral Risk Score Surveillance System (BRFSS) (https://www.cdc.gov/brfss/), a cross-sectional telephone survey that state health departments conduct monthly with a standardized questionnaire and technical and methodological assistance from CDC. BRFSS is used to collect prevalence data among adult residents in the United States regarding their risk behaviors and preventive health practices that can affect their health status. Respondent data are forwarded to CDC to be aggregated for each state. which reports self-reported adult obesity prevalence by race, ethnicity, and location (methodological details at http://www.cdc.gov/brfss/factsheets/pdf/DBS_BRFSS_survey.pdf). Data can be found at: https://data.cdc.gov/Behavioral-Risk-Factors/BRFSS-Table-of-Overweight-and-Obesity-BMI-/fqb7-mgjf

### Nighttime Light Exposure Data Acquisition

Nighttime light exposure data was generated by the www.lightpollutionmap.info platform (Jurij Stare, www.lightpollutionmap.info) which maps radiance data from NASA’s VIIRS/NPP Lunar BRDF-Adjusted Nighttime Lights Yearly composites (AllAngle_Composite_Snow_Free).

The Day/Night Band (DNB) sensor of the Visible Infrared Imaging Radiometer Suite (VIRRS), on board the Suomi-National Polar-orbiting Partnership (S-NPP) and Joint Polar Satellite System (JPSS) satellite platforms, provide daily measurements of nocturnal visible and near-infrared (NIR) light. NASA developed a suite of products of nighttime lights (NTL) applications including NASA’s Black Marble product suite (VNP46/VJ146). NASA’s Black Marble nighttime lights product, at 15 arc-second spatial resolution, is available from January 2012-present with data from the VIIRS DNB sensor. The VNP46/VJ146 product suite includes the daily at-sensor top of atmosphere (TOA) nighttime lights (NTL) product (VNP46A1/VJ146A1), daily moonlight-adjusted nighttime lights product (VNP46A2/VJ146A2), monthly moonlight-adjusted nighttime lights product (VNP46A3/VJ146A3), and yearly moonlight-adjusted nighttime lights product (VNP46A4/VJ146A4). To remove residual background noise, NASA’s VIIRS/NPP Lunar BRDF-Adjusted NTL composite values with radiances less than 0.5 nW.cm-2.sr-1 are set to zero. The retrieval algorithm, developed and implemented for routine global processing at NASA’s Land Science Investigator-led Processing System (SIPS), utilizes all high-quality, cloud-free, atmospheric-, terrain-, vegetation-, snow-, lunar-, and stray light-corrected radiance to estimate daily nighttime lights and other intrinsic surface optical properties. Details about algorithms, operational processing, evaluation, and validation are found in the user guide (Roman 2021). Data for each year include from January 1^st^ through December 31.

VIRRS data were overlaid on a map and areas of interest (i.e., states, counties) were outlined in triplicate (three independent measurements) and values were averaged to develop the average nighttime light intensity for each area of interest. Radiance data are provided as 10^−9^ W / cm^2^ * sr. World Atlas images were developed using “Falchi, Fabio; Cinzano, Pierantonio; Duriscoe, Dan; Kyba, Christopher C. M.; Elvidge, Christopher D.; Baugh, Kimberly; Portnov, Boris; Rybnikova, Nataliya A.; Furgoni, Riccardo (2016): Supplement to: The New World Atlas of Artificial Night Sky Brightness. GFZ Data Services. http://doi.org/10.5880/GFZ.1.4.2016.001 Falchi F, Cinzano P, Duriscoe D, Kyba CC, Elvidge CD, Baugh K, Portnov BA, Rybnikova NA, Furgoni R. The new world atlas of artificial night sky brightness. Science Advances. 2016 Jun 1;2(6):e1600377.”

### Statistics

Average nighttime light intensity data were generated for each state (excluding Alaska and Hawaii) or county and data were analyzed as an average of 2012-2018 as well as each year independently for each state. States were divided into five groups and county data divided into four groups according to average nighttime light intensity and an analysis of variance (ANOVA) was conducted followed by a post hoc Tukey for pairwise comparisons (GraphPad Prism 10.0.2) to compare AD prevalence among the groups. Correlation analysis was also conducted with state and county data to evaluate the relationship between AD prevalence and average nighttime light exposure (GraphPad Prism 10.0.2).

A linear mixed model was then applied to examine the relationship with all data together taking account of within-state or within-county correlation due to repeated measures. The state and county assessments included age, sex, and race in the model. For the state data, biological covariates (atrial fibrillation, chronic kidney disease, depression, diabetes, heart failure, hyperlipidemia, hypertension, obesity, and stroke) were considered and subgroup analyses were performed including: age (65+, <65), sex (men, women), and race (Asian Pacific Islander, Hispanic, Native American, Non-Hispanic Black, Non-Hispanic White). Analyses were conducted using SAS (v 9.4).

### Study Approval

The de-identified data presented in this report were obtained from public sources and do not constitute human subject research and has been deemed exempt by the Institutional Review Board at Rush University Medical Center.

## Data Availability

All data are available in the main text or the supplementary materials.

## Author Contributions

Conceptualization, Methodology, Investigation, Visualization, Writing – original draft, Writing, Editing, Funding: RMV; Methodology, Investigation, Visualization, Writing, Editing: BO, Writing, Funding: AK

---

**Fig. S1:**
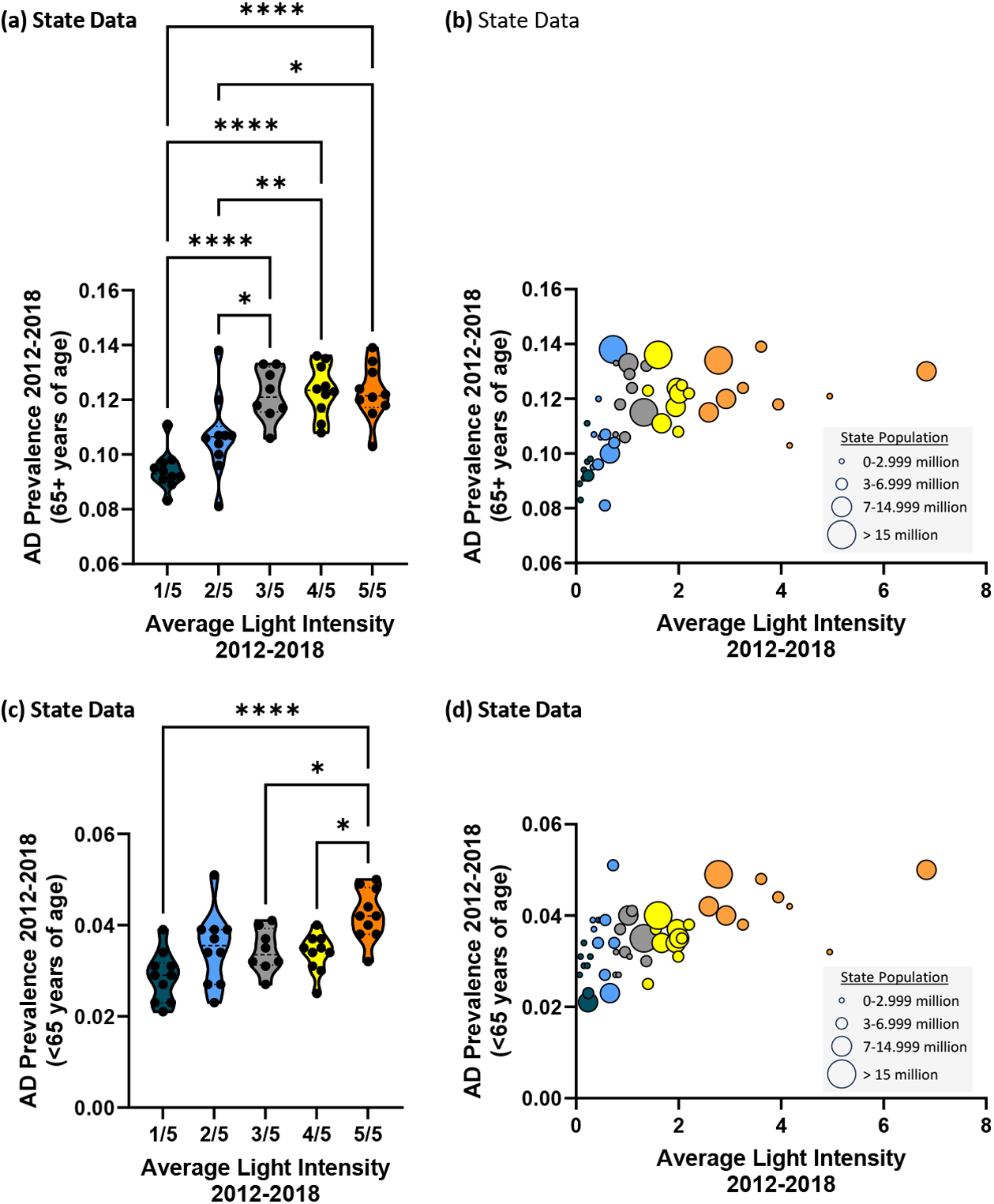
Higher state average nighttime light intensity is associated with higher state AD prevalence (2012-2018) in those over the age of 65 and those under the age of 65. **(a)** AD prevalence by state average nighttime light intensity rankings in individuals over the age of 65. ANOVA: F(4,43)=14.560, p<0.001. Multiple comparisons testing revealed differences between groups with lower average light intensity and groups with higher average light intensity: 1/5 vs. 3/5 (p<0.001), 1/5 vs. 4/5 (p<0.001), 1/5 vs. 5/5 (p<0.001), 2/5 vs. 3/5 (p=0.031), 2/5 vs. 4/5 (p=0.009), 2/5 vs. 5/5 (p=0.013). **(b)** Pearson correlation analysis between AD prevalence and nighttime light intensity in those over the age of 65: r(46)=0.490, p<0.001. **(c)** AD prevalence by state average nighttime light intensity rankings in individuals under the age of 65. ANOVA: F(4,43)=6.950, p<0.001. Multiple comparisons testing revealed differences between groups with lower average light intensity and groups with higher average light intensity: 1/5 vs. 5/5 (p<0.001), 3/5 vs. 5/5 (p=0.048), 4/5 vs 5/5 (p=0.018). **(d)** Pearson correlation analysis between AD prevalence and nighttime light intensity in those over the age of 65: r(46)=0.550, p<0.001. *p<0.05, **p<0.01, ***p<0.001, ****p<0.0001. Circle size reflects state population.

**Table S1:**
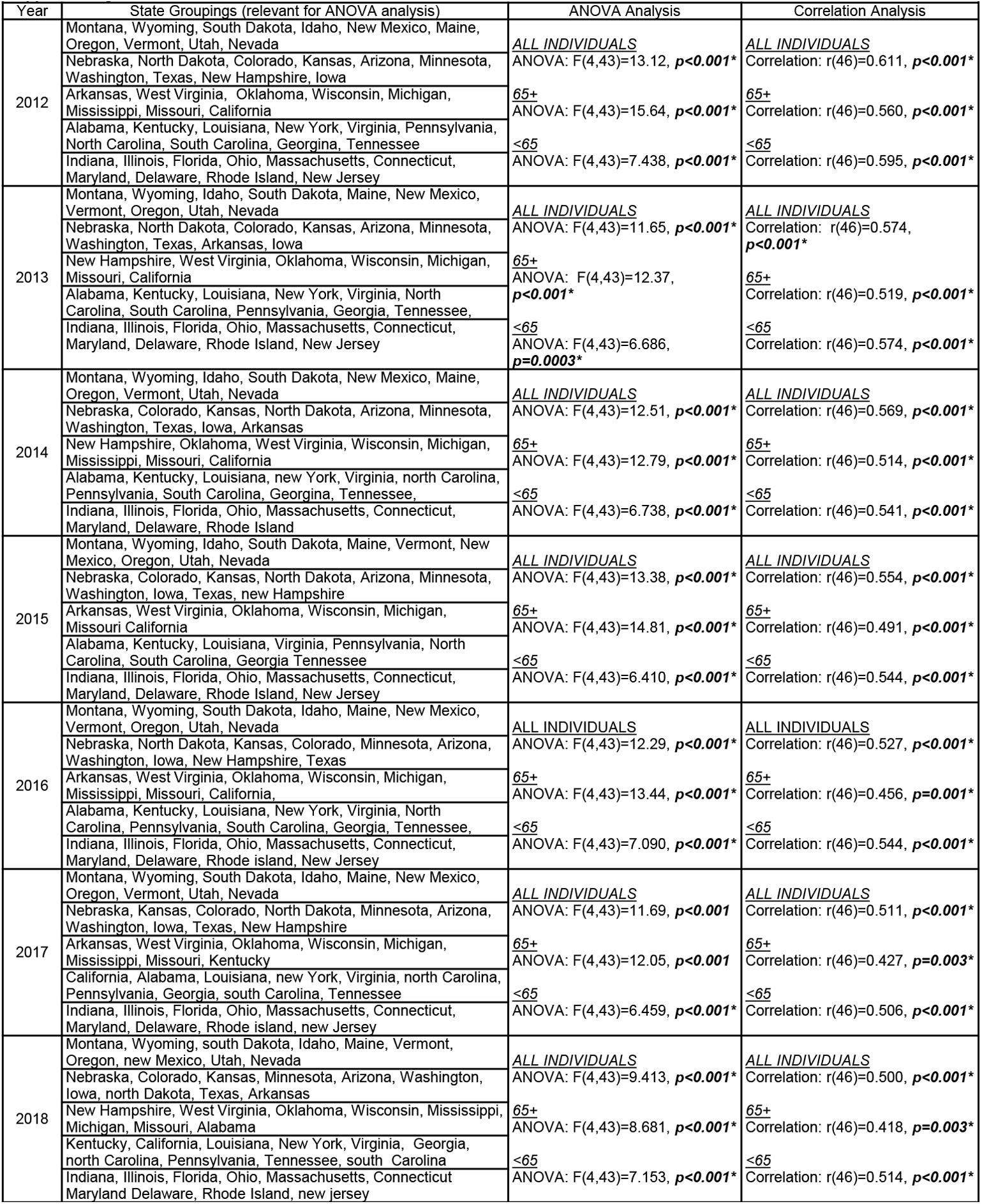
Analysis of State AD prevalence and average nighttime light intensity from 2012-2018. Each year (2012-2018) was individually assessed for the relationship between AD prevalence and average nighttime light intensity. A positive relationship was observed between AD prevalence and nighttime light intensity for each year. States were grouped according to highest to lowest average nighttime light intensity and these data were analyzed by analysis of variance (ANOVA). Results indicate a significant relationship between AD prevalence and light pollution). Linear correlation analysis was conducted between AD prevalence and average nighttime light intensity which demonstrated a significant positive correlation.

**Fig. S2:**
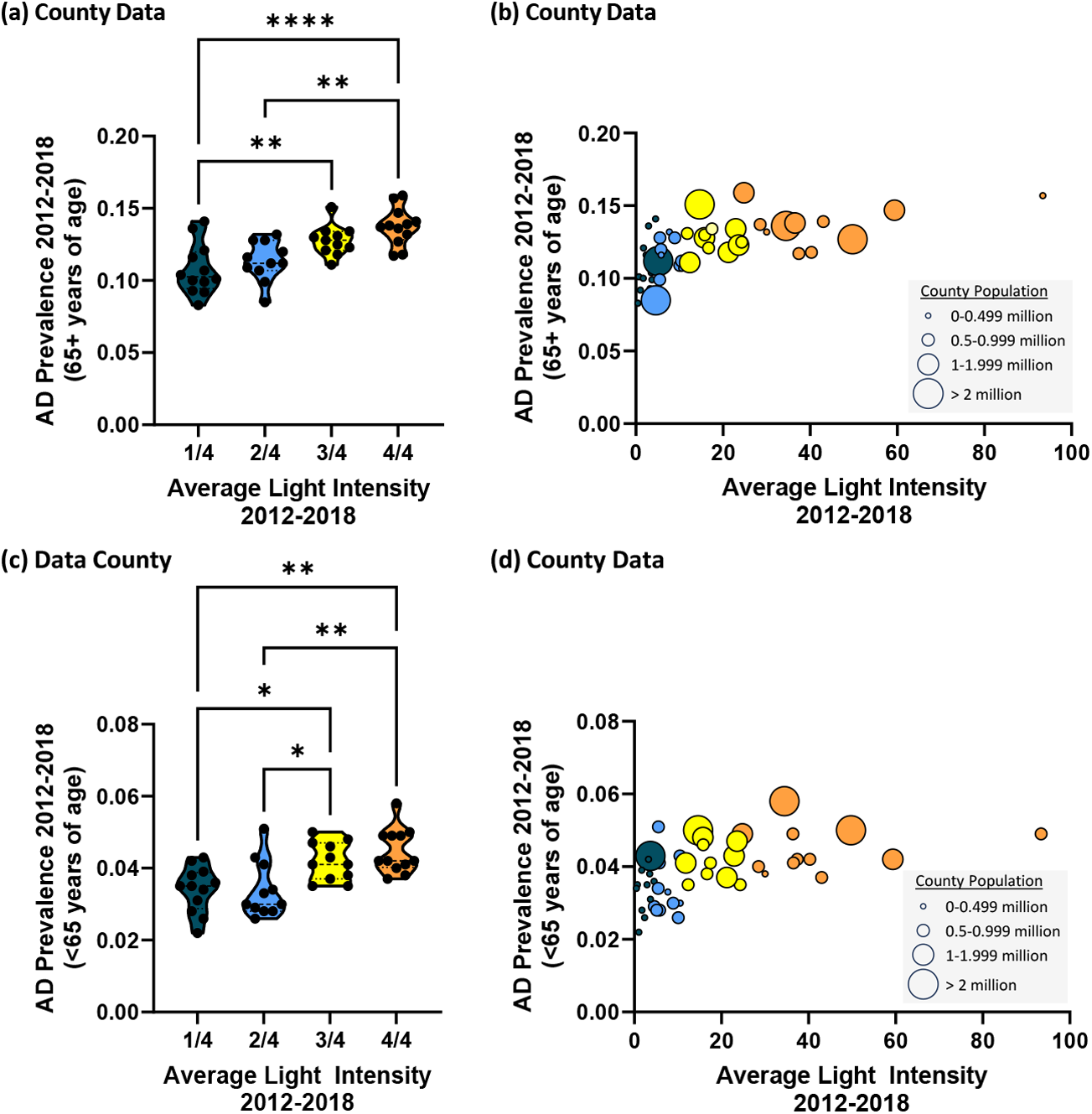
Higher county average nighttime light intensity is associated with higher county AD prevalence (2012-2018) in those over the age of 65 and those under the age of 65. **(a)** AD prevalence by county average nighttime light intensity rankings in individuals over the age of 65. ANOVA: F(3,42)=10.84, p<0.001. Multiple comparisons testing revealed differences between groups with lower average light intensity and groups with higher average light intensity: 1/4 vs. 3/4 (p=0.007), 1/4 vs. 4/4 (p<0.001), 2/4 vs. 4/4 (p=0.001). **(b)** Pearson correlation analysis between AD prevalence and nighttime light intensity in those over the age of 65 (r(44)=0.612, p<0.001). **(c)** AD prevalence by state average nighttime light intensity rankings in individuals under the age of 65. ANOVA: F(3,42)=8.424, p<0.001. Multiple comparisons testing revealed differences between groups with lower average light intensity and groups with higher average light intensity: 1/4 vs. 3/4 (p=0.030), 1/4 vs. 4/4 (p=0.001), 2/4 vs 3/4 (p=0.029), 2/4 vs. 4/4 (p=0.001). **(d)** Pearson correlation analysis between AD prevalence and nighttime light intensity in those over the age of 65 (r(44)=0.514, p<0.001. *p<0.05, **p<0.01, ****p<0.0001. Circle size reflects county population.

**Fig. S3:**
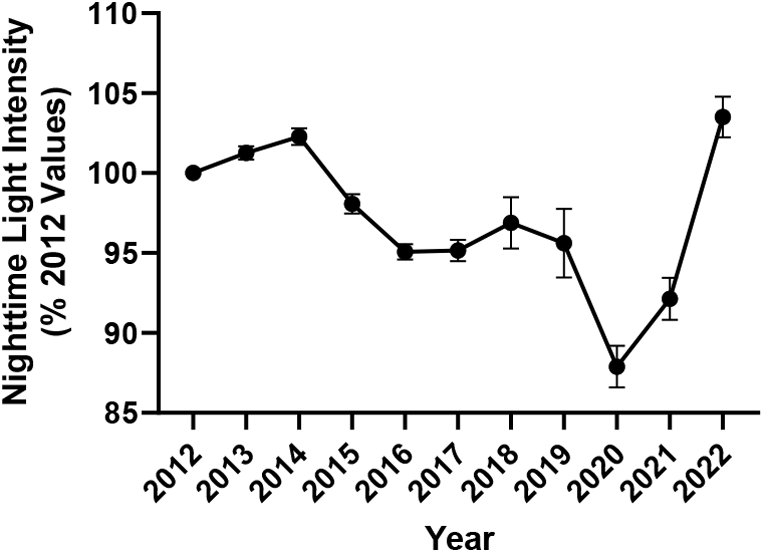
Changes in nighttime light from 2012-2022 at the state level. Nighttime light in 2012 (for each state) was set as 100% and changes in light across years 2013-2022 was calculated.

